# Estimating the basic reproduction number at the beginning of an outbreak under incomplete data

**DOI:** 10.1101/2021.07.14.21260514

**Authors:** Sawitree Boonpatcharanon, Jane Heffernan, Hanna Jankowski

## Abstract

We compare different methods of estimating the basic reproduction number, *R*_0_, focusing on the early stages of an epidemic, and considering weekly reports of new infecteds. We study three standard epidemiological models: SIR, SEIR, and SEAIR and examine the sensitivity of the estimators to the model structure. As some methods are developed assuming specific epidemiological models, our work adds a study of their performance in both the well- and miss-specified settings. We focus on parameters matching various types of respiratory viruses, although the general approach is easily extendable to other scenarios.

## 1 Introduction

The basic reproduction number, *R*_0_, (also called the basic reproductive ratio) is defined as the expected number of new infections produced by a single (typical) infectious individual, when introduced into a totally susceptible population. *R*_0_ is used in epidemiological studies of infectious diseases to gauge how contagious/transmissible an infectious disease is: if *R*_0_ < 1, the disease will die out, and if *R*_0_ > 1 infection can increase in the population. It is also used to determine how effective vaccination or other disease mitigation strategies need to be in order to protect populations from infection.

At the outset of an infectious disease outbreak, an immediate goal is to determine *R*_0_, so that public health and healthcare decision makers can be informed. At the debut of the COVID-19 pandemic, reports of *R*_0_ estimates were plentiful (for examples, see Zhao et al. (2020); Tuite and Fisman (2020); Knight and Mishra (2020); Mellan et al. (2020); Hilton and Keeling (2020); Price et al. (2020)). In the recent MERS-COV, 2009 H1N1, and 2003 SARS epidemics, there were also numerous studies of *R*_0_ globally (see Nishiura et al. (2010); Chowell et al. (2011); Tuite et al. (2010); Paine et al. (2010); Fraser et al. (2009); Pourbohloul et al. (2009); Chowell et al. (2014); Hsieh (2015); Cauchemez et al. (2014); Riley et al. (2003); Anderson et al. (2004); Wang and Ruan (2004); Dye and Gay (2003) for a small snapshot).

There are many statistical and mathematical methods that can be used to estimate *R*_0_ (Heffernan et al., 2005; Diekmann et al., 1990, 2010; van den Driessche and Watmough, 2002; Vegvari et al., 2021; Heesterbeek and Dietz, 1996; Blumberg and Lloyd-Smith, 2013; Gallagher et al., 2020; Farrington et al., 2001; White et al., 2021). Typical studies of *R*_0_ will thus employ a comparison of several estimators to provide increased certainty in *R*_0_ values. Different estimators also, however, can be constructed on different assumptions related to disease characteristics i.e., serial interval, infectious period, and thus may ignore the effects of different stages of infection. For example, many *R*_0_ estimators have been constructed to work within a Susceptible-Infectious-Recovered (SIR) disease modelling framework. Infectious diseases, however, can include periods of infection that are not infectious. The infectious period can also be split into various stages of asymptomatic and symptomatic infection, which ultimately affect the case reporting rate to public health. Therefore, methods that are based on the SIR modelling framework can project erroneous estimates of *R*_0_, and differences in *R*_0_ estimates may simply reflect poor estimator structure or application to data that has been misspecified. These aspects make it difficult to compare *R*_0_ estimates to gain increased certainty.

A recent study by Gallagher et al. (2020) has discussed several nuances of different estimator methods that can affect *R*_0_ estimates. The effect of data misspecification is only touched on briefly. Herewithin, we provide a detailed analysis of misspecified data using six different *R*_0_ estimators. The current study is organized as follows. We first provide an introduction to three compartmental infectious disease models that we use to generate data. Six *R*_0_ estimators are then introduced, including a discussion of their underlying compartmental model structure assumptions. We then apply each estimator to data generated from the three compartmental models. We employ parameter values representative of respiratory virus epidemics, and in particular, influenza Cowling et al. (2009); Vink et al. (2014); Park and Ryu (2018). We note that while daily data may be sometimes available during an infectious disease outbreak, it may not be complete and can include a reporting delay. We thus have chosen to use weekly case reports. Weekly case report data is also typical to outbreaks of influenza, a respiratory virus, and our chosen pathogen of study.

## 2 Methods

### 2.1 Epidemiological models

We focus on three compartmental epidemiological models, the Susceptible-Infectious-Recovered (SIR), Susceptible-Exposed-Infectious-Recovered (SEIR), and Susceptible-Exposed-Asymptomatic Infectious-Symptomatic Infectious-Recovered (SEAIR) models (model equations are provided in the Appendix A.1). All models are considered without inclusion of demography, i.e. birth and death. The total population is denoted by *N* with initial values of *S*_0_ and *I*_0_ for *S* and *I* populations, respectively, such that *N* is approximately equal to *S*_0_. That is, for the SIR model, for all *t ≥* 0 it holds that *S*(*t*) + *I*(*t*) + *R*(*t*) = *S*_0_ + *I*_0_ = *N* . Similarly, *S*(*t*) + *E*(*t*) + *I*(*t*) + *R*(*t*) = *N* for the SEIR model and for the SEAIR model, *S*(*t*) + *E*(*t*) + *A*(*t*) + *I*(*t*) + *R*(*t*) = *N* . We use the notation *θ* = (*β, σ, ρ, γ*) to denote the vector of parameters for the models, see Table 1. For each model, the associated formulas for *R*_0_ and the serial distribution are listed in Table 2.

**Table 1:** Contact rate notation in epidemiological models

| model | parameter |  |  |  |
| --- | --- | --- | --- | --- |
| | $\beta$ | $\sigma$ | $\rho$ | $\gamma$ |
| SIR | $S \rightarrow I : \beta I(t)/N$ | | | $I \rightarrow R : \gamma$ |
| SEIR | $S \rightarrow E : \beta I(t)/N$ | $E \rightarrow I : \sigma$ | | $I \rightarrow R : \gamma$ |
| SEAIR | $S \rightarrow E : \beta I(t)/N$ | $E \rightarrow A : \sigma$ | $A \rightarrow I : \rho$ | $I \rightarrow R : \gamma$ |

**Table 2:** Summary of epidemiological model properties. To obtain the serial distribution, let *Y*_1_, *Y*_2_, *Y*_3_ be independent exponential random variables with mean one.

| model | $\theta$ | $R_0 = R_0(\theta)$ | serial distribution |
| --- | --- | --- | --- |
| SIR | $(\beta, \gamma)$ | $\beta/\gamma$ | $Y_1/\gamma$ |
| SEIR | $(\beta, \gamma, \sigma)$ | $\beta/\gamma$ | $Y_1/\gamma + Y_2/\sigma$ |
| SEAIR | $(\beta, \gamma, \sigma, \rho)$ | $\beta/\gamma + \beta/\rho$ | $Y_1/\gamma + Y_2/\sigma + Y_3/\rho$ |

Data is generated using the SIR, SEIR, and SEAIR compartmental model structures using a stochastic agent-based modelling framework implemented in C++. The simulations progress at the level of individual hosts in the applicable model disease status compartments. The simulation moves forward using “event times” that are assigned to each infected individual in the population, whereby the event times correspond to the disease model compartment of which they are currently a member. Such event times correspond to infection events, when an infected individual transmits the infection to a susceptible, and times at which infected individuals progress to the next stage of infection or recover. The C++ model is based on previous work Heffernan and Wahl (2005, 2006). 1000 simulations are conducted for each of the SIR, SEIR, and SEAIR frameworks with parameters (*β, σ, ρ, γ*) = (5*/*9, 1, 1, 1*/*3), giving *R*_0_ values of 5/3 for the SIR and SEIR models, and 20/9 for the SEAIR model (see Table 2 for formulas). For each epidemic, the population size *N* is set to 10, 001 where *S*(0) = 10, 000 and *I* (0) = 1.

Figure 1 plots the number of individuals in compartment *I* for each model structure. The grey lines plot the simulation outcomes while the black lines plot the mean of the simulation data. Although the complete epidemic path is simulated, we assume that only the weekly number of infectious people is actually available. The epidemics are followed for 15 weeks, which covers the first 100 days of an outbreak. Simulation data is recorded at every event time. Weekly data is extracted from each simulation and saved in a data file for use for all of the *R*_0_ estimators employed here. The blue vertical line indicates the point of inflection, where the concavity of the black line changes. The inflection points were observed at 7, 12, and 9 weeks respectively for the SIR, SEIR, and SEAIR models. These points are used to determine appropriate time intervals for *R*_0_ estimation for each model since *R*_0_ estimates are associated with early exponential growth and can be affected by decreases in the growth rate as the epidemic continues towards and past the point of infection.

**Figure 1:**
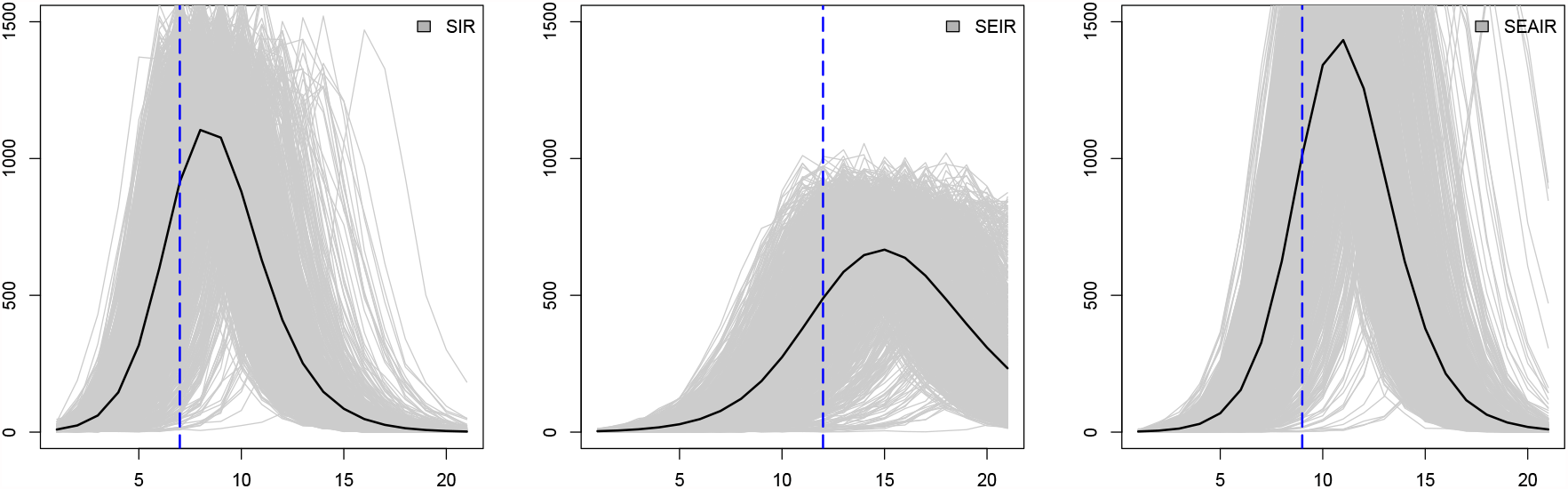
Plots of numbers of infectious individuals (*y*-axis) at time *t* in weeks (*x*-axis); from left to right: SIR, SEIR, and SEAIR. Individual simulated outbreaks from 1000 simulations are shown as grey lines, and their average is denoted as a black line. The blue vertical dashed lines show the inflection points for each model.

### 2.2 Estimating *R*_0_

Many methods exist to estimate *R*_0_, and we refer to White et al. (2021) for a recent review. If the transition rates in the models of Section 2.1 are known, then *R*_0_ can be easily calculated using the formulas listed in Table 2. However, full transition rates are generally not known in practice, and hence statistical estimation methods are required. The main difficulty in estimation is that complete epidemic data is unavailable. Here, we consider six different methods of estimating *R*_0_. These are detailed below. A summary of the methods and their key properties is also given in Table 3.

**Table 3:** Summary of estimation methods

| method |  |
| --- | --- |
| WP | serial distribution can be assumed known or can be estimated using MLE; method developed under branching process model; simple method which yields real-time estimates |
| secB | serial distribution assumed known (only the mean is used); method developed assuming SIR model; simple method which yields real-time estimates |
| ID/IDEA | serial distribution assumed known (only the mean is used); method developed assuming SIR model; simple method which yields real-time estimates |
| plug-and-play | serial distribution assumed unknown; method selects one of SIR/SEIR/SEAIR model; implementations available though not real-time (depending on input selection) |
| fullBayes | serial distribution assumed unknown; method selects one of SIR/SEIR/SEAIR model; not real-time |

The first four (WP, secB, ID, and IDEA) are real-time methods based on simplifications of the full ODE epidemiological models. This simplification is necessitated by the fact that the full data is unobservable. In these methods, estimation of *R*_0_ is coupled with either estimation or prior knowledge of the serial distribution. The serial distribution is the distribution of the random amount of time that an individual is infected. For example, in the SIR model, the serial distribution is exponential with mean 1*/γ* (see Table 2 for other models). In the literature, the serial distribution may also be referred to as the serial interval, although this most often refers to the mean of the serial distribution, or alternatively, a range indicating highly likely values from the serial distribution. As our focus here is the seasonal flu, it may be reasonable to assume that the serial distribution is known apriori. For other situations, such as new emerging diseases, such assumptions are less valid.

The two latter methods (plug-n-play and fullBayes) do not simplify the full epidemic models, but handle the issue of unobservable data by Monte Carlo simulation (plug-n-play method) or Bayesian priors with MCMC used to handle estimation due to model complexity (fullBayes method). As such, these methods are more computationally intensive. These two methods estimate the unknown transition rate parameter vector *θ* in the epidemic model. They do not require any prior knowledge, including prior knowledge of the serial distribution. Indeed, since the methods result in estimates of *θ*, these can then in turn be used to derive an estimate of the serial distribution. Furthermore, the methods assume prior knowledge of the epidemic model, in the sense that the user can decide whether the SIR, SEIR, or the SEAIR model is more appropriate for the particular disease. In contrast, the first four methods all rely on simplifications, and are not able to allow for such tailoring.

Although the last two methods are more computationally intensive and not considered “real-time”, we note that modern day access to computational power is blurring this line of distinction. Our implementations of fullBayes and plug-n-play were done on a non-specialized desktop computer and without special consideration to computing time in the implementations. The time required to obtain the estimates was less than two minutes in both cases, and we do not consider this to be prohibitive. Furthermore, more careful programming could yield even faster estimates. A more detailed discussion is available in Section 3.1.

#### 2.2.1 Maximum likelihood estimation of a branching model (WP method)

White and Pagano (2008) developed a straightforward estimation method whereby either the distribution of the serial interval is known, or, the distribution of the serial interval is estimated along with *R*_0_. The method assumes that only the number of infectious individuals at discrete time points (e.g. daily or weekly) is observable. Both methods rely on maximum likelihood. Using our notation, and assuming that the times *t*_0_ = 0, *t*_1_, *t*_2_, …, *t*_*k*_ are integers which count, for example, the number of days or weeks, White and Pagano (2008) obtain the log-likelihood

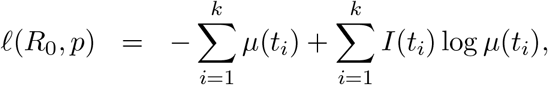

where 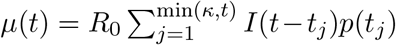 and *p* is a vector denoting the distribution of the serial interval on *t*_1_, …, *t*_*κ*_. If *p* is assumed known (notably, this includes knowing the value of *t*_*κ*_ which describes the support of *p*) then the maximum likelihood estimate of *R*_0_ is straight-forward to compute. For example, in the SIR model, *p*(*t*_*j*_) = *P* (*t*_*j*−1_ < *Y ≤ t*_*j*_)*/P* (*Y ≤ t*_*κ*_) where *Y* is an exponential random variable with mean 1*/γ*. If *p* is unknown, then White and Pagano (2008) recommend assuming a parametric distribution to simplify estimation.

The WP method assumes an underlying branching process, which is neither of the SIR/-SEIR/SEAIR models from which our data sets are generated. This model assumes, in particular, that throughout the population size “available” to be infected remains constant, which does not hold for our simulated ODE models. As such, estimates should only really be considered early on in the epidemic. In our simulations presented below, we highlight the inflection point of each epidemic, and the WP method should only really be considered valid before this time.

The method has been implemented in Obadia and Boëlle (2015), see also Obadia et al. (2017). In our simulations, we found this implementation to have some numerical instability issues, which is most likely caused by the particular parameters of our simulated data sets. This instability was particularly profound when *p* was assumed unknown, and most often the algorithm would not yield a solution. For this reason, we programmed our own implementation, for which we used a simple grid search. The built-in alternative optimization function in R uses the bisection method, and was very sensitive to the starting value (a small change in the starting value could change the *R*_0_ estimate by orders of a thousand). In comparison, the grid search approach performed better, although it was still not ideal. The likelihood surface is very flat, which resulted in a non-unique MLE (we report only a default value). This property of the likelihood surface is most likely what also causes the issues in the implementation of Obadia et al. (2017).

Although not reported with our main results (which use the gamma assumption suggested by White and Pagano (2008)), we also tried other approaches to estimating the SD distribution. The easiest to implement (via a one-dimensional grid search) is the assumption that the serial distribution is any distribution with a support of at most two weeks. In Figure 2 we compare this assumption with the assumption that the serial distribution is gamma but still unknown. We also compare this with two cases where the serial distribution is known. Here, the serial distribution is exponential with either a mean of five days or a mean of three days. The true serial distribution under the SIR model is exponential with a mean of five days in our simulations, so the second setting is misspecified. Due to the weekly nature of the data and resulting discretization on the serial distribution in the WP method, the effect of this misspecification is very mild.

**Figure 2:**
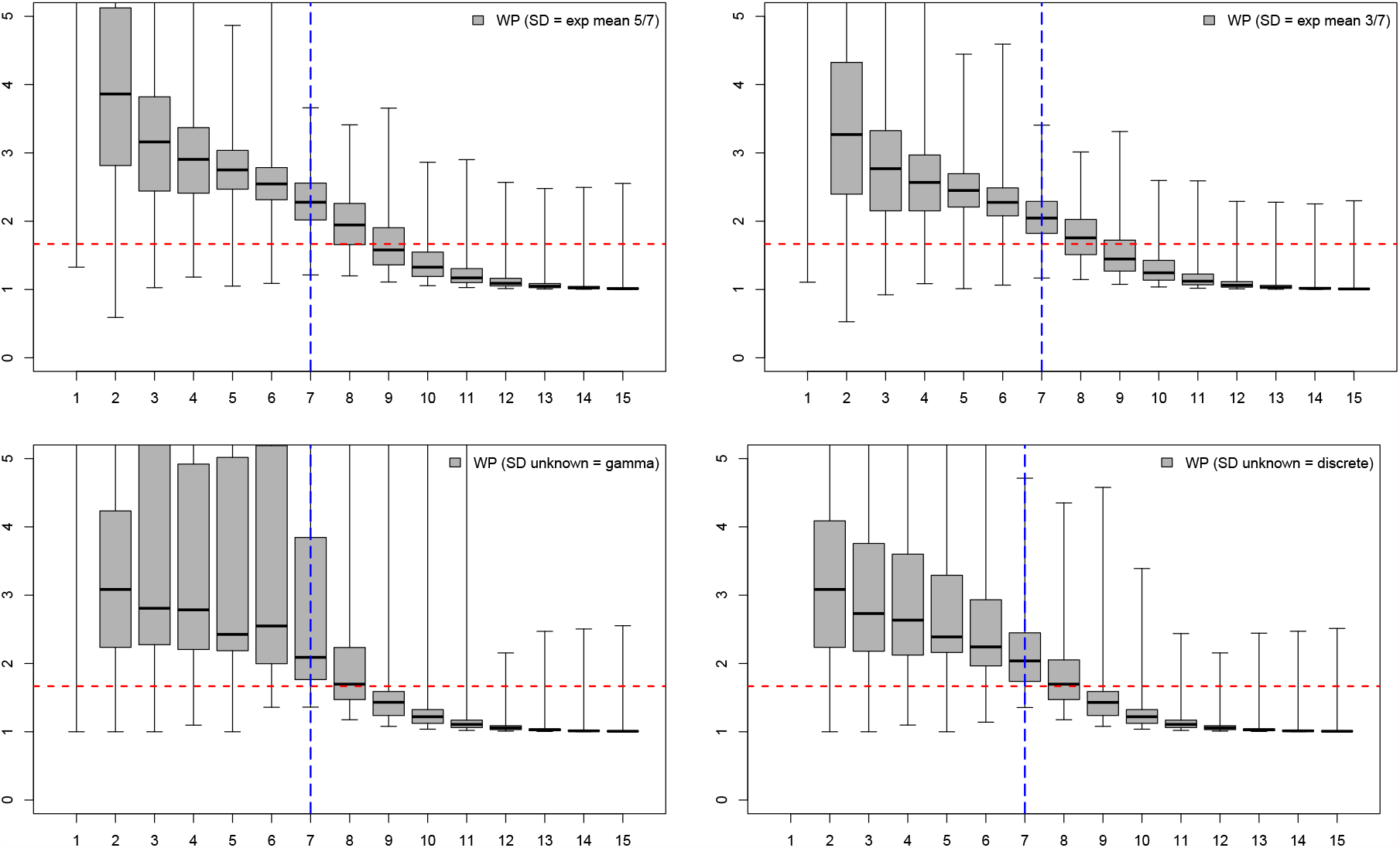
WP method for SIR data with four different assumptions on the serial distribution: known and correct (top left), known and incorrect (top right), unknown gamma (bottom left), unknown discrete (bottom right). The inflection point for the epidemic is marked in blue, and the true *R*_0_ is marked as a horizontal red line.

Furthermore, note that the log-likelihood assumes that the serial distribution is discrete, and that this discretization matches the observed data. That is, if data is observed weekly, the serial distribution is only known *on a weekly timescale*. This discretization can affect the serial distribution considerably, particularly if the timescale is quite coarse. We also note that the implementation of Obadia and Boëlle (2015) automates the discretization, and therefore we suggest that care is taken when using their built-in parametric distribution functions.

#### 2.2.2 Sequential Bayes estimation using an SIR approximation (secB method)

Bettencourt and Riberio (2008) developed a Bayesian approach used to estimate *R*_0_. As above, it is assumed that infectious counts are observed at periodic times such as days or weeks. The basic idea is to start with a mildly informative prior on *R*_0_ and then update sequentially. The approach is based on the SIR model, and assumes that the mean of the serial distribution is known (under the SIR model, this is equivalent to knowing the parameter *γ* which is the inverse of the mean of the serial distribution). Bettencourt and Riberio (2008) note that under the SIR model

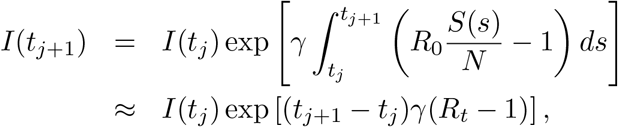

where *R*_*t*_ = *R*_0_*S*(*t*)*/N* ≈ *R*_0_ at the beginning of an infection. Using this result, seqB assumes that the conditional distribution of *I*(*t*_*j*+1_)|*I*(*t*_*j*_), *R*_0_ is Poisson with mean *λ* = *I*(*t*_*j*_) exp{(*t*_*j*+1_ − *t*_*j*_)*γ*(*R*_0_ − 1)}. In the approach, *γ* is known, and a prior is placed on *R*_0_. With *N*_0_ also assumed known, posterior estimates are found using a hierarchical or sequential Bayes approach. Note that the method cannot handle data sets where there are no new infections observed in some time interval *t*_*j*+1_ − *t*_*j*_ (as this results in a Poisson mean of zero). Therefore, the times at which infectious counts are observed must be sufficiently coarse so that all counts are non-zero (e.g. weeks instead of days). The method would also be inappropriate for situations where long intervals between cases are observed in the initial stages of the epidemic. This was observed, for example, in Canada for the first cases of Covid19.

Although the above development is based on the SIR model, the resulting approximation behaves similarly to a branching process, much like the WP method. We therefore again consider this estimator valid only in the early stages, which for our simulations translates to times prior to the inflection points of the epidemic.

The posterior distribution of *R*_0_ will have the same support as the prior, and placing a discretized prior on *R*_0_ makes computations relatively straightforward, since the normalizing constant of the posterior is easy to implement. In the R implementation in Obadia et al. (2017), the initial prior on *R*_0_ is assumed to be uninformative. Their package focuses on the posterior mode, and much like their implementation of the WP method, uses a discretized version of the serial distribution (which could affect the input value of *γ*). We again chose to use our own implementation, and report the posterior mean which minimizes the Bayes’ risk.

#### 2.2.3 Least square estimation using incidence decay approximations (ID and IDEA methods)

Fisman et al. (2013) introduced two simplified models describing the relationship between *R*_0_ and other epidemic parameters in the SIR model. The first of these is the incidence decay (ID) model where

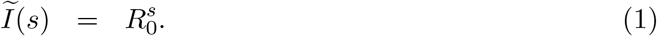

In the model, time is measured in units re-scaled based on the serial distribution. Recall that under the SIR model the serial distribution is exponential with mean 1*/γ*. We then have the relationship in (1) that Ĩ(*s*) = *I*(*γs*). As (1) is only valid for a short (and unknown) period of time, Fisman et al. (2013) proposed a second alternative formulation, where a decay factor was introduced in order to reflect the often observed outbreak decline. In the incidence decay and exponential adjustment (IDEA) model, the relationship becomes instead

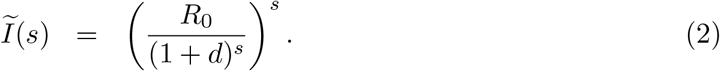

Under the ID model, we can solve (1) to obtain

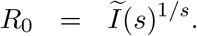

Of course, this relationship is not valid for real data across all values of *s* as Ĩ(*s*) is stochastic. To obtain an estimate of *R*_0_ least squares is an obvious option, and hence the ID estimator is the minimizer of

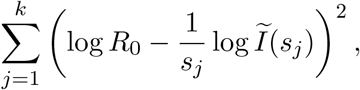

which yields

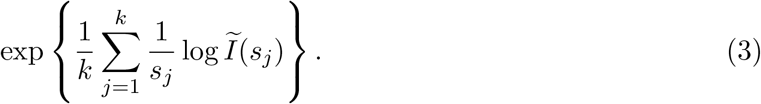

As noted above, the number of infectious people decreases rapidly at the beginning of an outbreak, so a method based on (1) is expected to underestimate *R*_0_. The IDEA model was introduced to overcome this issue. As in the ID model, we solve (2)

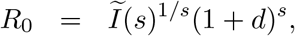

and use least squares estimation to obtain its estimate. The IDEA estimator is defined then as the minimizer of

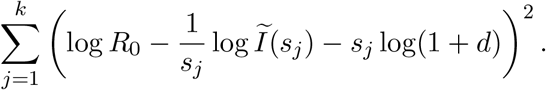

Unlike in the ID model, we also need to obtain a minimizer of *d* to solve the optimization problem, and hence we require *k ≥* 2. Minimizing, we obtain

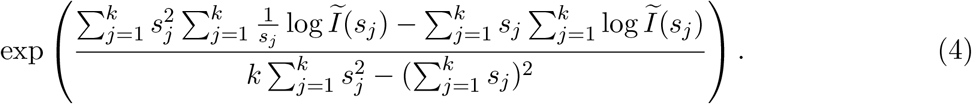

Details of these calculations are given in the Appendix. Note that the formula is not valid for *k* = 1.

Both the ID and IDEA methods are straightforward and estimate *R*_0_ directly, as long as the mean of the serial distribution is known. The model was built under the SIR assumption, however. In our simulations we examine the effect of misspecification of the underlying epidemic model.

#### 2.2.4 Maximum likelihood using sequential Monte Carlo for partially observed epidemics (plug-n-play method)

Maximum likelihood is one of the more popular approaches used to estimate unknown parameters in a statistical model. The general idea is to find the parameter set *θ* which maximizes the likelihood (probability model) evaluated at the observed data. The difficulty for our setting is that our epidemiological models (Section A.1) rely on data which is unobservable. In particular, the models require that the exact times of infections are known while we observe only daily or weekly counts of infectious individuals. The WP method (White and Pagano, 2008), which also uses maximum likelihood, gets around this issue by creating a simplified model with a likelihood which relies only on observable data. Another alternative, discussed in He et al. (2010), is to maximize the full likelihood and fill in the unobservables using many Monte Carlo simulations in a way which matches the fixed observable data points. Such an approach is often referred to as “plug-n-play”.

The plug-n-play inferential method of He et al. (2010) is based on likelihood inference using sequential Monte Carlo of partially observed Markov processes (POMP), also known as hidden Markov models or state-space models. The plug-and-play terminology comes from the fact that inference is based on Monte Carlo simulations from the model and does not require explicit expressions of the transition probabilities, which can be quite complicated. The algorithm for this method has been implemented in Nguyen et al. (2016). This software package can be accessed from the comprehensive R archive network (CRAN), see King et al. (2017). As mentioned previously, the basic idea is to generate complete epidemic data in a way which matches the observed weekly infectious observations. To simplify the implementation, complete continuous-time data is not generated but rather an approximation is generated with observations of all components at a discretized time-scale Δ*t* (selected by the user). These discretized epidemics are generated using sequential Monte Carlo methods. An estimate of *θ* is then obtained via maximum likelihood using iterated filtering. The implementation in Nguyen et al. (2016) allows for the selection of the model SIR, SEIR, or SEAIR. The algorithm requires initial values for the rate parameters *θ*, the number of ‘particles’ *J* used in the sequential Monte Carlo method, choice of the time scale Δ*t*, while the iterated filtering requires some further choice of algorithm settings. We refer to He et al. (2010) and Nguyen et al. (2016) for additional details. The algorithm returns estimates of *θ*, as well as an estimate of *R*_0_ derived via the formula

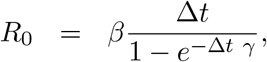

regardless of the epidemiological model. We refer to the estimate thus obtained as the plugn-play estimator. R code detailing our simulations and choices of input values is given in Appendix A.2.

#### 2.2.5 Bayesian inference for partially observed epidemics (fullBayes method)

Similarly to the plug-n-play approach of the previous section, this is a simulation approach in which the incomplete observed data is replaced with complete data via simulations. The main difference is that the complete data is generated by placing a prior on its distribution in a Bayesian inferential approach. Some examples of epidemiological inference under the Bayesian paradigm are described in O’Neill and Roberts (1999).

In order to describe the method we need first to introduce some additional notation. We do this for the SEAIR model, as all other models are simplifications of this case. Recall that we have observed infection counts *I*(*t*_1_), …, *I*(*t*_*k*_) at times *t*_1_, …, *t*_*k*_. Let *m* denote the vector with *j*th element given by 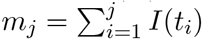. As such, *m* describes the entirety of the observed data. For a time interval [0, *T*] the complete epidemic includes much more information. Let 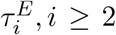 denote the individual times of exposure. Similarly, 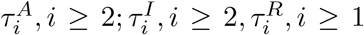 denote the individual times of transitions into the asymptomatic, infectious, and recovered states, respectively. We assume that *m*_0_ = 1. We also assume that all people who are infected in week *j* will recover in week *j* + 1. Furthermore, we assume that the number of exposed and asymptomatic people in week *j* is also equal to *m*_*j*_ − *m*_*j*−1_. We let *τ* denote the epidemic path which contains all of this information.

As in O’Neill and Roberts (1999), the first infection 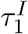 is treated separately as a parameter of the model. Hence a prior 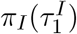 is placed on this variable. An independent prior is also placed on *θ, π*(*θ*), and samples from the posterior distribution 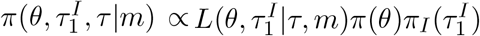 are obtained. The marginal distribution of 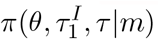, which is the posterior distribution of *θ* given the observable data, and the distribution we are interested in.

We now calculate the likelihood 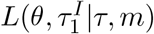 for the SEAIR model.

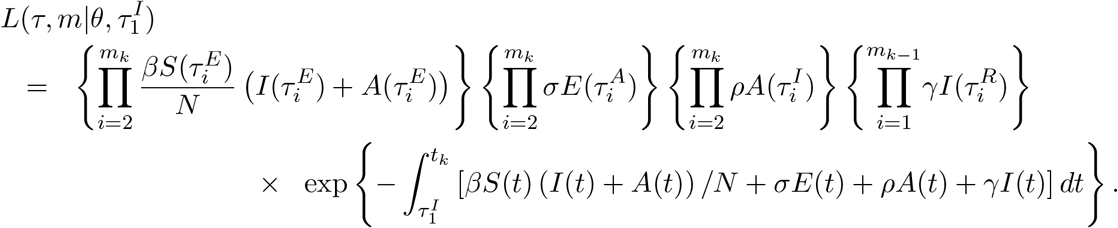

The joint prior distribution of the unknown rate parameters *θ* is made up of independent gamma distributions given by Γ(*α, k*) with mean *k/α*. We assume that *α* is the same for the parameters *β, σ, ρ, γ*, while *k* varies and if appropriate will be denoted by *k*_*β*_, *k*_*σ*_, *k*_*ρ*_, *k*_*γ*_. In the simulations we take *α* = 1 and *k*_*β*_ = *k*_*σ*_ = 3, *k*_*ρ*_ = 2, *k*_*γ*_ = 5. The prior distribution on 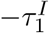 is exponential with rate one, and this is independent from the *θ* vector. Calculations given in the Appendix (see Section A.4) give the posterior marginal distributions for 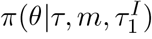 and 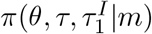 all of which have gamma distribution with closed form expressions for the parameters. Some sensitivity analysis to the prior distributions was conducted in Section A.6, and changing the prior did not visibly affect the results.

The general approach we take is now described using the following steps.

1. Use Markov chain Monte Carlo (MCMC) to simulate from 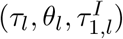.
2. From Step 1, we obtain a sequence of samples 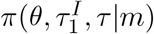 for *l* = 1, …, *b* + *B* from the posterior distribution 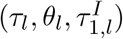. Here, *b* denotes the burn-in period for the MCMC results. To obtain an estimate of *θ*, from the samples *l* = *b* + 1, *b* + *B*, one option is to simply average the values *θ*_*l*_. Instead, we treat each 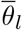 a sample from the full posterior model, and calculate the posterior mean of *θ*_*l*_, using the formulas given in the Appendix.
3. Average the posterior means 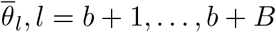, *l* = *b* + 1, …, *b* + *B* to obtain an estimate of *θ*.

The final reported estimate is obtained from the estimate of *θ* in Step 3 using the appropriate formula in Table 2. In our simulations, we take *b* = 100 and *B* = 1000, and refer to the estimator as fullBayes.

The MCMC algorithm we use is the Metropolis-within-Gibbs. Namely, there are three main components to the posterior distribution *θ, τ*, and 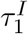. In the Appendix, the posterior distributions for 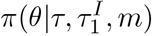 and 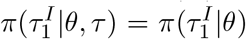 are obtain in closed form. Given one observation of 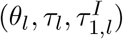, the algorithm generates the next observation as follows.

1. Sample 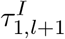 from the posterior 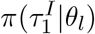.
2. Sample *θ*_*l*+1_ from the posterior 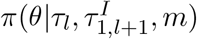
3. Sample *τ*_*l*+1_ using a Metropolis step:
  a. Propose a new *τ* : For each *i* = 1, …, *k*
    i. 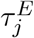 is IID uniformly distributed on [*t*_*i*−1_, *t*_*i*_] for *j* = *m*_*i*−1_, …, *m*_*i*_
    ii. 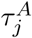 is IID uniformly distributed on [*t*_*i*−1_, *t*_*i*_] for *j* = *m*_*i*−1_, …, *m*_*i*_
    iii. 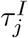 is IID uniformly distributed on [*t*_*i*−1_, *t*_*i*_] for *j* = *m*_*i*−1_, …, *m*_*i*_
    iv. 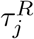 is IID uniformly distributed on [*t*_*i*−1_, *t*_*i*_] for *j* = *m*_*i*−2_, …, *m*_*i*−1_
  b. Accept the proposal with probability min{1, *α*} where

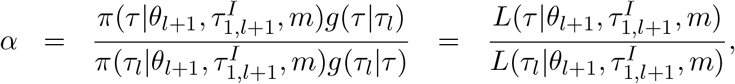

noting that with the proposal distribution in (a), we have that *g*(*τ* |*τ*_*l*_)*/g*(*τ*_*l*_|*τ*) = 1. Details are provided in Appendix A.4

The chain is initialized by sampling *θ* from its prior distribution.

## 3 Results

The goal of our simulations is to study accuracy of estimation in the well-specified but also in the misspecified settings, including misspecification of the model and serial distribution. For all models we therefore consider data coming from SIR, SEIR, and SEAIR settings. We then study the methods as follows

1. WP method assuming
  - SD is known and set to exponential with mean of 5 days (correct assumption under SIR model)
  - SD is unknown and estimated from a gamma distribution with unknown mean and variance (using a grid search algorithm)
2. seqB method assuming
  - SD has a mean of 5 days (i.e. *γ* = 7*/*5)
  - SD has a mean of 3 days
3. ID and IDEA methods assuming
  - SD has a mean of 5 days (i.e. the serial interval is 5*/*7 in the language of Fisman et al. (2013))
  - SD has a mean of 3 days
4. plug-n-play and fullBayes methods developed assuming
  - SIR
  - SEIR
  - SEAIR

In our set-up, the outbreaks were all followed for 15 weeks, and this is the timeline given in our results. This timeline is presented only as a comparison to what is happening at the earliest stages. It also, however, improves the comparison between methods. Our comments below focus only on the time period before the inflection point (denoted as a vertical blue line for all methods).

Side-by-side boxplots summarizing our simulation results are given in Figures 3-8. Recall that the seqB method cannot handle data sets where zero values are present. We have thus removed simulations with zero values (3 SIR, 41 SEIR, and 76 SEAIR epidemics) from the 1000 samples for seqB method. They are included for the other methods. In comparing the efficacy of the methods, we look at two main factors: bias and variability.

**Figure 3:**
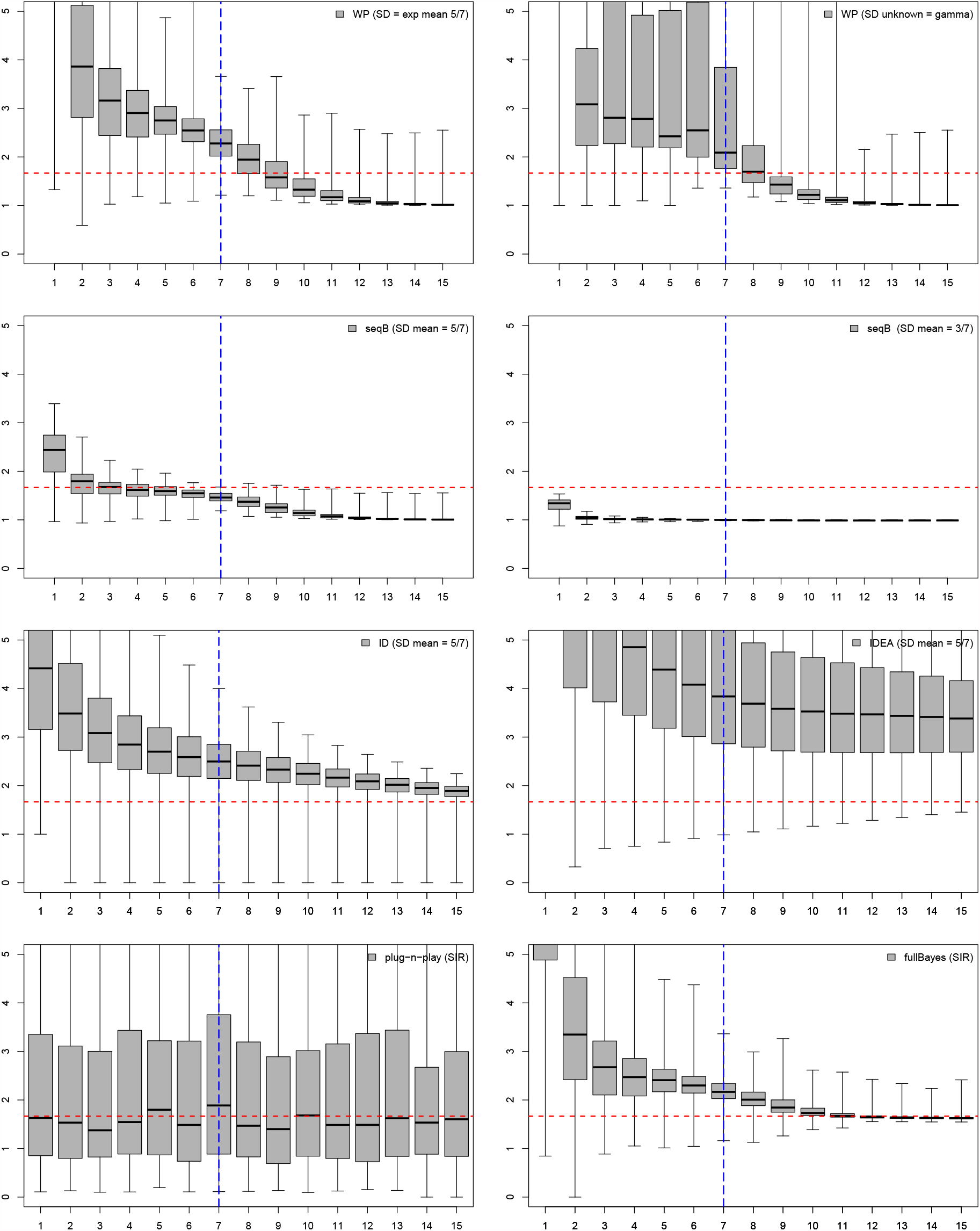
*R*_0_ estimates assuming SIR model with SIR data (week on *x*-axis). True *R*_0_ is given by the red dashed horizontal line, with the inflection point indicated by the blue dashed vertical line.

Figures 3 and 4 study the methods assuming that the correct model assumption is used. In Figure 3 we also include the seqB method with incorrect serial distribution for reference. Considering both bias and variance, of the methods with known SD, seqB performs best when consider the SIR model (Figure 3, second row, left column). For the SEIR and SEAIR models (Figure 4), and of the methods with unknown SD for the SIR model (Figure 3), fullBayes seems best. Although plug-n-play has small bias, in our view this is overshadowed by the extremely large variability of the method. In general, these observations carry forward into our misspecification studies.

**Figure 4:**
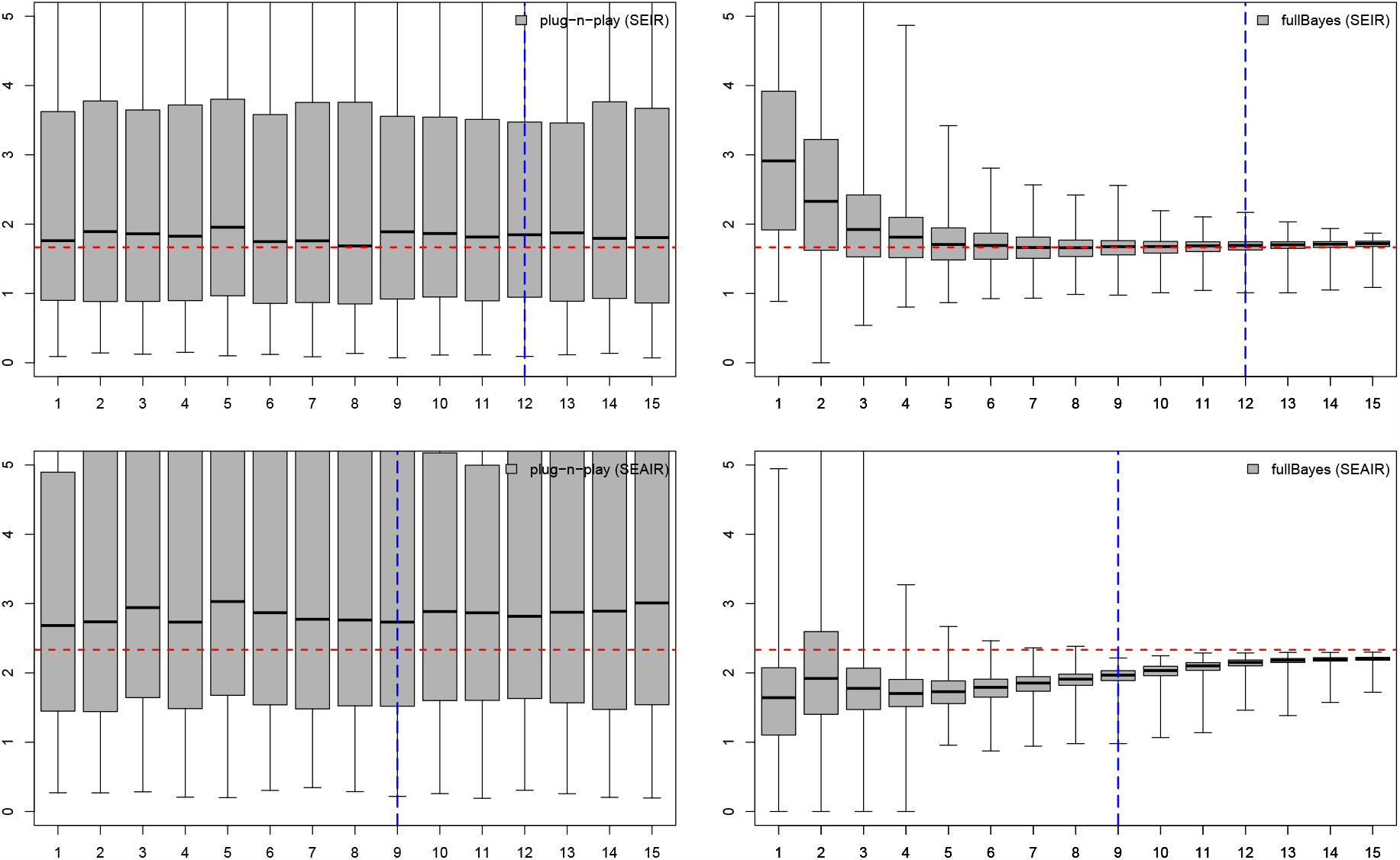
*R*_0_ estimates assuming SEIR model with SEIR data in the top row and SEAIR model with SEAIR data in the bottom row (week on *x*-axis). True *R*_0_ is given by the red dashed horizontal line, with the inflection point indicated by the blue dashed vertical line.

Figure 5 focuses on methods which assume SIR and known SD, and considers misspecification of the serial distribution. Note that here the mean of the serial distribution was incorrect by only two days. Notably, in comparing the left and right columns of this figure, WP does not appear very sensitive to a change to incorrect mean, while the other methods are more sensitive.

**Figure 5:**
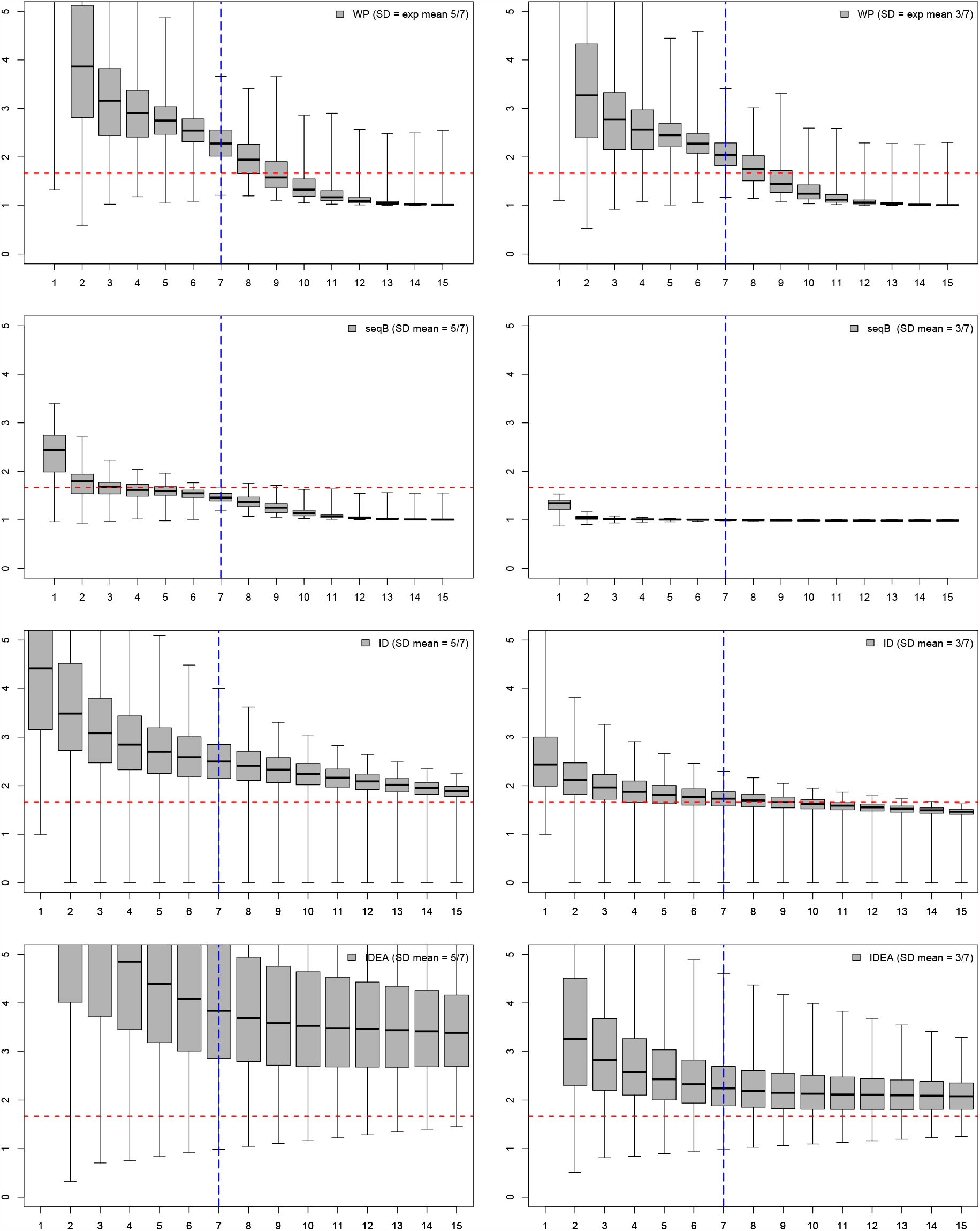
*R*_0_ estimates assuming SIR model with SIR data (week on *x*-axis) for methods which assume SD is known with correct (left column) and incorrect (right column) assumptions on SD. True *R*_0_ is given by the red dashed horizontal line, with the inflection point indicated by the blue dashed vertical line.

Figures 6, 7 and 8 study model misspecification. In Figure 6 the model assumed is SIR while the data is SEIR. In Figure 7 the model assumed is SIR while the data is SEAIR. Finally, in Figure 8 the model is SEIR while the data is SEAIR. Note that only plug-n-play and fullBayes can assume the SEIR model. In Figures 6 and 7, WP with known SD performs well. With unknown SD, the estimate quality decreases in both bias and variability. In all cases fullBayes when the serial distribution is unknown performs very well.

**Figure 6:**
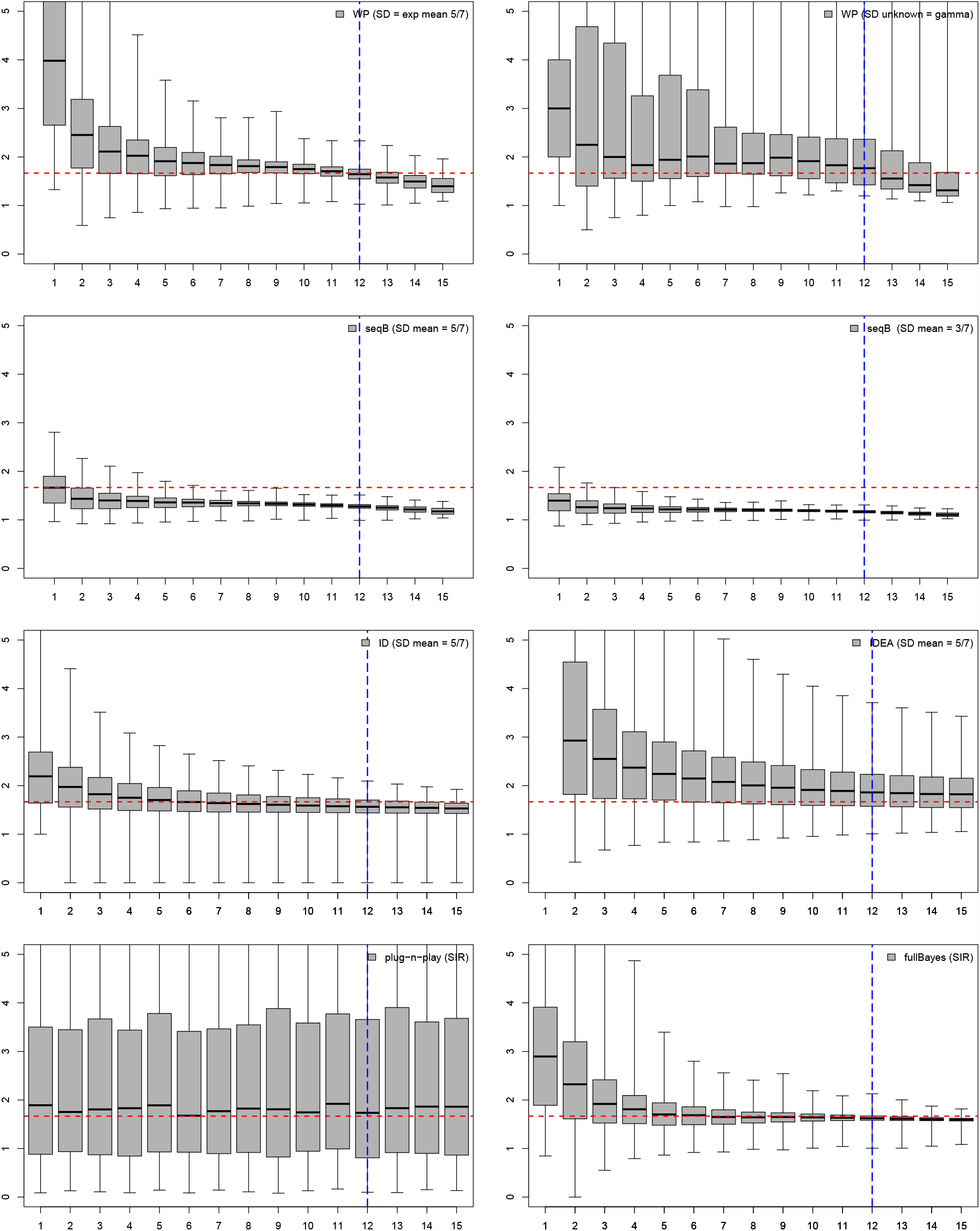
*R*_0_ estimates assuming SIR model with SEIR data (week on *x*-axis). True *R*_0_ is given by the red dashed horizontal line, with the inflection point indicated by the blue dashed vertical line.

**Figure 7:**
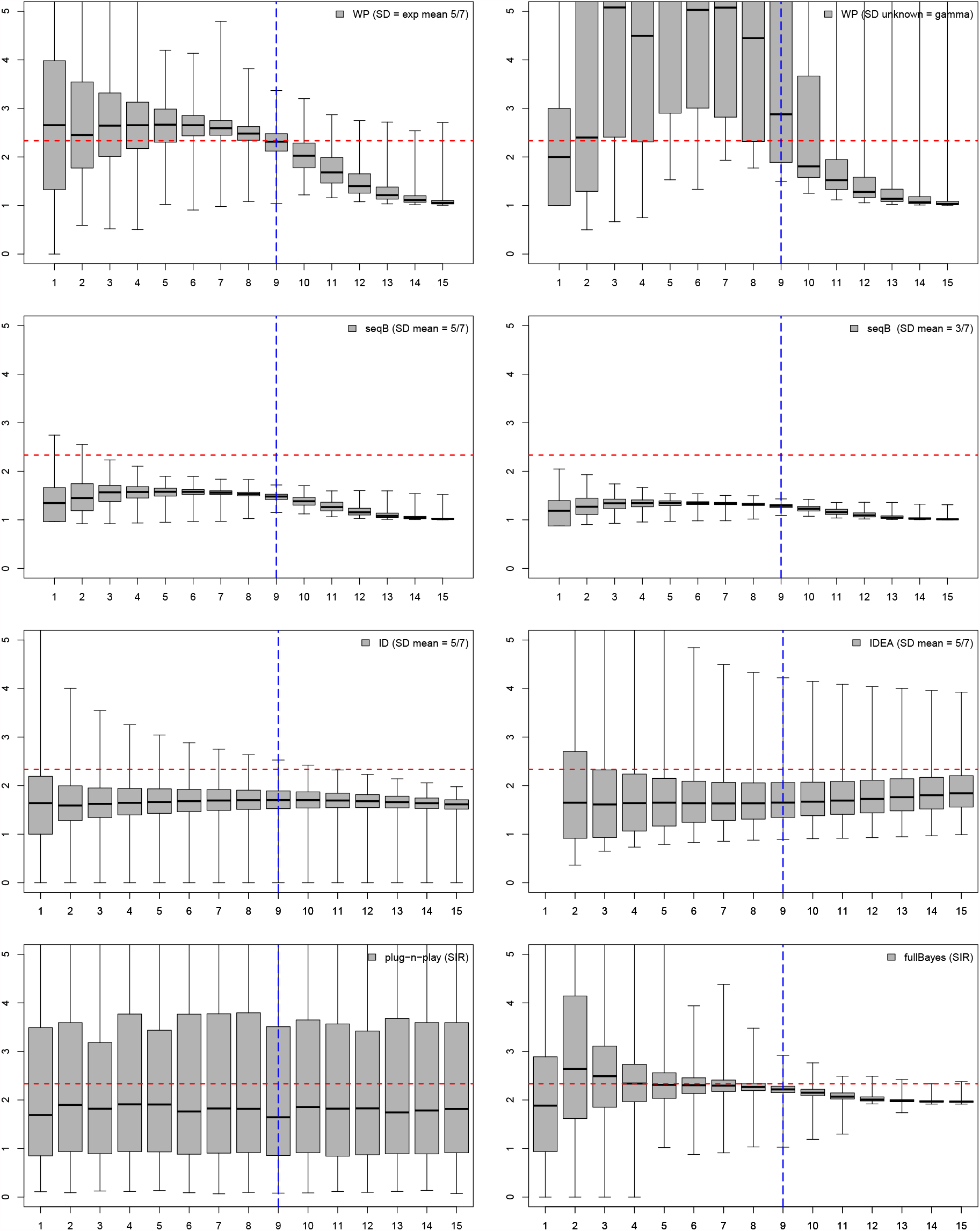
*R*_0_ estimates assuming SIR model with SEAIR data (week on *x*-axis). True *R*_0_ is given by the red dashed horizontal line, with the inflection point indicated by the blue dashed vertical line.

**Figure 8:**
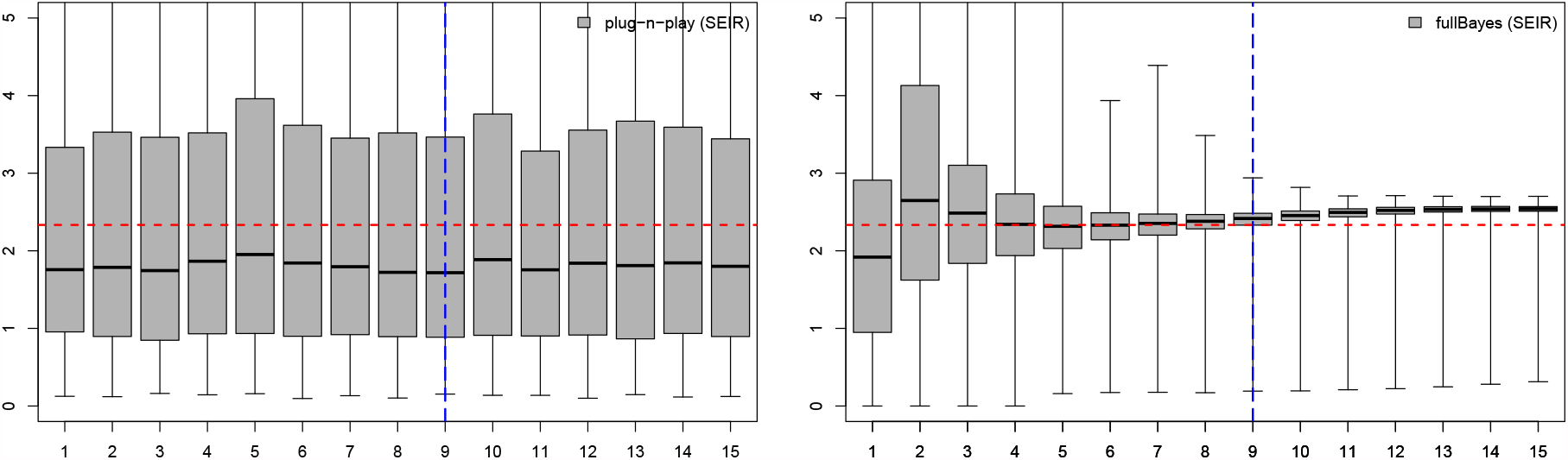
*R*_0_ estimates assuming SEIR model with SEAIR data (week on *x*-axis). True *R*_0_ is given by the red dashed horizontal line, with the inflection point indicated by the blue dashed vertical line.

Based on the simulations, our recommendations are as follows. In general, we recommend the fullBayes method assuming SIR, unless another model (e.g. SEIR, SEAIR) is preferred. Although the method is not considered to be real-time, we feel the computational burden is not overly onerous as estimates will be given in 1-2 minutes, depending on the computer’s power and speed. In particular, fullBayes performs well even under some misspecification of the empidemiological model, and does not require prior knowledge of the serial distribution. The alternative option here is the WP method, which did not perform as well under the gamma assumption for the serial distribution in our simulation study. Alternative assumptions (such as the discrete distribution assumed in Figure 2) may also be considered, and numerical stability should be checked before applying these algorithms. If the serial distribution is known and the epidemiological model is SIR, we recommend the seqB method, but emphasize the need for sensitivity analysis.

Practitioners, however, should consider their own preferences as to bias and variability of the estimators. We note here that as this study is focused on data observed weekly, our results may not be applicable to data observed, for example, daily, as the effect of the serial distribution on the results may be different.

### 3.1 Computational time

Computational time is a crucial factor as real-time estimates are desirable. Table 4 shows computational time for the SEIR model for a single data set and using a 1.60GHz/8GB RAM desktop PC. The results in this work are based on fullBayes with 1000 iterations and plug-n-play with 1000 particles and 10 IF iterations, where IF stands for the iterated filtering algorithm. The fullBayes method was implemented in R, and it is possible that faster implementations can be achieved using a different programming language.

**Table 4:**
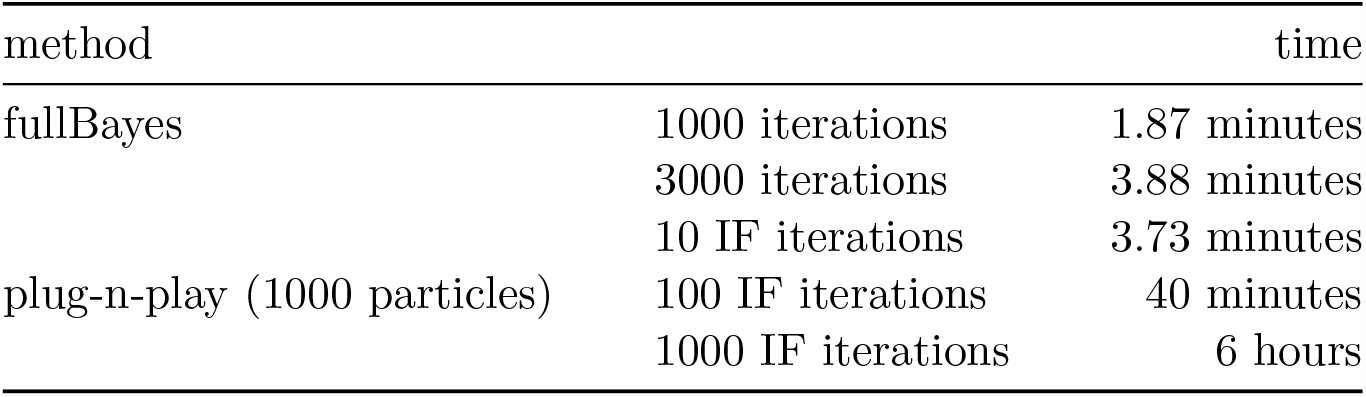
Computational time for the SEIR model for one data set

## 4 Conclusion

The basic reproduction number, *R*_0_, is an important parameter for estimation early in an epidemic so that public health interventions can be informed. As many estimators exist, and the assumptions of the estimators as well as their dependency on particular biological estimates i.e., the serial interval, vary between methods, it is expected that *R*_0_ estimates will differ. It is thus important to understand what estimators provide better outcomes under both true and misspecified conditions. Since respiratory viruses (especially influenza) affect the global population every year, we have chosen to study the estimators of *R*_0_ for these types of infections, which are typically modelled using SIR, SEIR and SEAIR compartmental models. We have also chosen to consider weekly case data, as this is characteristic of influenza and other respiratory infection outbreak reported data, globally.

We have considered six estimators that are commonly used when determining *R*_0_ for any infectious disease outbreak. The advantages and disadvantages of each method are discussed here, including dependencies on proper estimates of the serial distribution, and the computational resources needed to run each estimator. Briefly, we find that the WP method can provide close estimates to the true *R*_0_ value if the SD is known, but that the method suffers numerical instability in cases when the SD is unknown for the type of data considered here; the seqB method performs well given SIR data but greatly underperforms if there is any misspecification; the ID and IDEA methods, although useful for other purposes due to their simplicity, do not outperform any of the methods studied here in terms of estimating *R*_0_; the plug-n-play method estimates include large confidence intervals, so do not provide precise *R*_0_ estimates; the fullBayes method is the least sensitive to model structure and misspecification. Considering both bias and variability, as well as misspecification, we find that the performance of the fullBayes estimator is best, providing estimates of *R*_0_ that are closest to the true value under both correctly specified and misspecified cases. The approach also does not require prior knowledge of the serial distributions. We note that the choice of *R*_0_ estimator is ultimately up to the practitioner. Our strong recommendation, however, is that if simpler methods are chosen, a full sensitivity analysis considering the misspecifications studied here, should be employed so that confidence in an *R*_0_ estimate can be acquired.

In our analysis we have shown that some *R*_0_ estimators can be greatly affected by even a small level of misspecification. Given that biological certainty may be lacking at the beginning of an infectious disease outbreak, the number of disease stages needed in a model and a proper distribution of the serial interval may not be known. This means that a range of *R*_0_ results will ensue, and the accuracy of the estimates will be unclear. We therefore recommend that the fullBayes method be included in any suite of estimators used to estimate *R*_0_ as it does not require knowledge of the serial distribution and provides close to true estimates under different model structures quickly.

Daily case reporting data has been available for the most recent COVID-19 pandemic. Daily data was not provided during the 2009 H1N1 pandemic, however. Furthermore, there may be issues with daily reporting (such as periodicity, reporting delay) whereby public health may choose to use weekly reporting data over daily data as the weekly data would be more reliable. We have thus only considered weekly case reporting data in this study as it is expected that weekly case reporting data can be expected in many future epidemics and pandemics. It is important to note that First Few Hundred (FF100) studies, whereby the first few hundred cases of a new virus are followed in detail at the beginning of an infectious disease outbreak, have been implemented during the 2009 H1N1 and COVID-19 pandemics Black et al. (2017); World Health Organization and others (2020); McLean et al. (2010); Boddington et al. (2020); van Gageldonk-Lafeber et al. (2012); England (2009); Ghani et al. (2009); Pandemic Influenza (2014). In these cases the serial distribution, and the need to consider exposed and/or asymptomatic periods of infection can be quickly determined, enabling realization of earlier and more certain estimates of *R*_0_ early on. Given that First Few Hundred protocols are not implemented in much of the globe, weekly case report data however may still be considered the norm for future pandemics.

In our current study we have assumed perfect data with no unobserved infections, no reporting delay, and no data collection bias. These issues are intuitively expected to affect *R*_0_ estimates. We venture to continue our study of *R*_0_ estimation while considering these aspects in our epidemiological data sets.

## Data Availability

We are developing a shiny app. The app will be available when the manuscript is accepted for publication.

## A Appendix

### A.1 Disease models

We employ SIR, SEIR and SEAIR model structures, where *S, E, A, I* and *R* denotes susceptible, exposed (infected, no symptoms, not infectious), asymptomatic infected (infected, no symptoms, not infectious), symptomatic infected (infected, symptomatic, infectious), and recovered individuals, respectively. The ODEs governing our models are given below.

#### A.1.1 SIR model

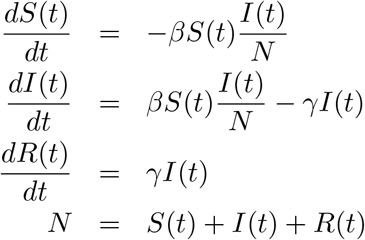

#### A.1.2 SEIR model

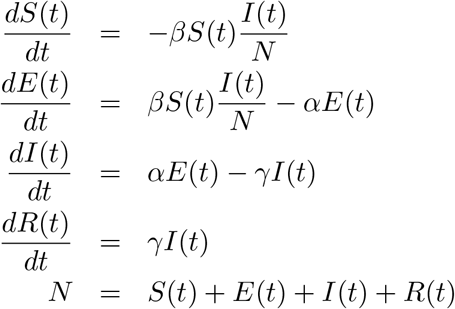

#### A.1.3 SEAIR model

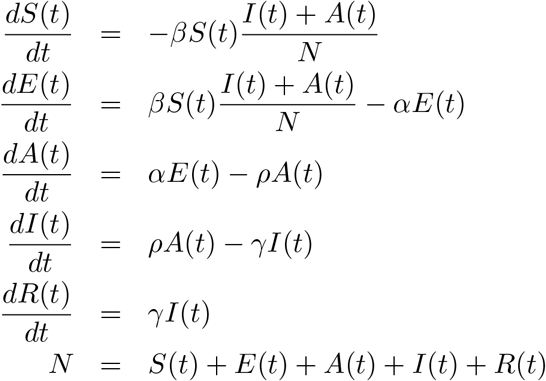

### A.2 Least squares estimation for the IDEA method

From (2), our objective function is

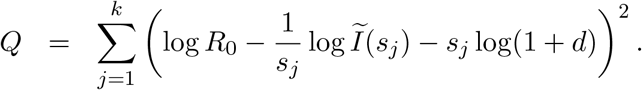

Let *η* = log *R*_0_ and ξ = log(1 + *d*) and note that both of these relationships are monotone increasing. We minimize *Q* by setting 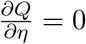 and 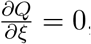, obtaining two equations

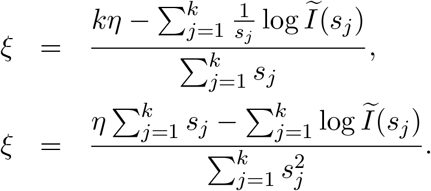

Solving for *η* = log *R*_0_ we thus find

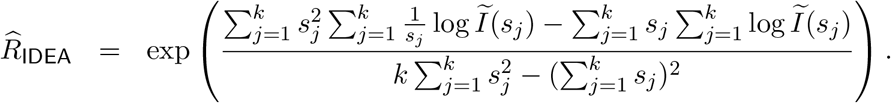

### A.3 R code to implement plug-n-play estimation for SIR model

~~~
l i b r a r y (pomp)
s e t . seed (1)
dat <− read . csv (“Weekly NewnumInf SIR . csv “)
dat <− dat [ 1, ]
dat <− as . numeric (dat)
dat <− cumsum(dat)
nc <− length (dat)
rho <− 1 ## all cases are reporte d
difft <− 1/ 7 ## dofiltering for every day
res beta <− rep (0, nc)
resgamma <− rep (0, nc)
R0e <− rep (0, nc)
R0 <− rep (0, nc)
   f o r (j in 1 : nc){
      data <− c (1, as . numeric (dat [ 1 : j ]))
      date <− as . numeric (seq (0, j, 1))
      infdat <− cbind (date, data)
      infdat <− as . data . frame (i n f d a t)
      colnames (i n f d a t) <− c (“week”, “cases“)
      ######## function ###########
      sir.proc.sim <− function (S, I, R, H, beta, gamma, delta . t, …){
      N <− sum(S, I, R)
      foi <− beta ∗ I /N
      trans <− c (reulermultinom (n=1, size=S, rate=f oi, dt=delta .      t),
       reulermultinom (n=1, s i z e=I, r a te=gamma, dt=d e l ta . t))
       S = S−trans [ 1 ]
       I = I+trans [ 1] − tr a n s [ 2 ]
       R = R+trans [ 2 ]
       H = H+trans [ 2 ]
       c (S=S, I=I, R=R, H=H)
}
f <− f u n c t i o n (t, S, I, R, beta, gamma){
      N <− sum(S, I, R)
      f o i <− beta ∗ I /N
      terms <− c (
          S∗ f o i,
          I ∗gamma
)
terms <− unname (terms)
c (
     S=−terms [ 1 ],
     I=terms [ 1] − terms [ 2 ],
     R=terms [ 2 ],
     H=terms [ 2 ]
  )
}
sir dmeas <− function (cases, H, rho, log, …) {
     #c a s e s = sum(infdat [, 2 ])
     dbinom (x=cases, size=H, prob=rho, log=log)
}
sir rmeas <− function (H, rho, …) {
     c a s e s=rbinom (n=1, size=H, prob=rho)
}
flu . sir <− pomp(data= i n f d a t,
                              times=“week”,
                              t 0 =0,
                              params=c (rho =1,gamma=mean (rgamma(n = 20, 1, 5)), beta=mean (rgamma(
                                         S. 0 =10000, R. 0 =0, I . 0 =1, H. 0 = 0),
                              rmeasure=s i r rmeas,
                              dmeasure=s i r dmeas,
                               rprocess=euler (sir . proc . sim, delta . t=d i f f t)
)
#################################### end f u n c t i o n ########################################
para <− coef (flu . sir)
simpar <− c (“beta”, “gamma”)
f i t <− mif 2 (f l u . s i r, Nmif=5,
                rw . sd=rw . sd (beta =0 . 01, gamma= 0 . 005),
             cooling . fraction . 50 = 0 . 01,
            Np=1000)
   suppress Warnings (f i t)
   resbeta [ j ] <− as . numeric (coef (fit) [ 3 ])
   resgamma [ j ] <− as . numeric (coef (fit) [ 2 ])
   IP <− d i f f t /(1−exp(− difft ∗resgamma [ j ]))
   R0e [ j ] <− resbeta [ j ] ∗IP
   R0 [ j ] <− resbeta [ j ] / resgamma [ j ]
}
~~~

### A.4 Posterior distributions

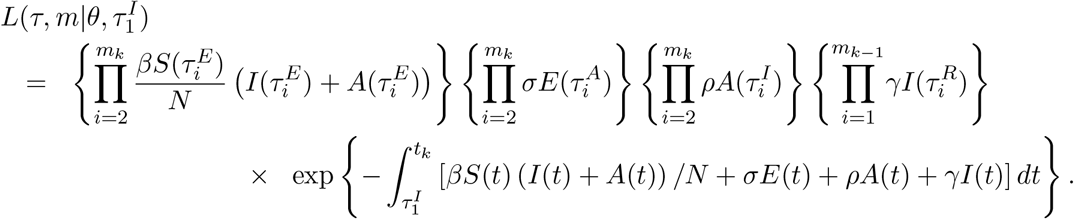

First, we calculate the posterior distributions of each of the elements of *θ*. For ease of exposition, in each calculation we drop the subscript on the scale parameter *k* in the priors.

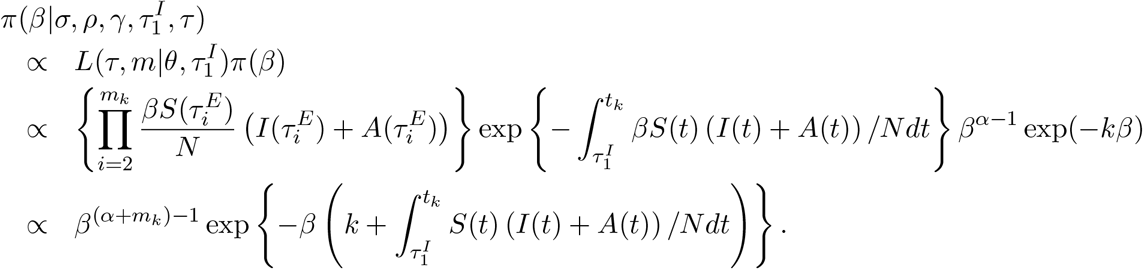

Hence the posterior distribution for *β* is gamma with shape parameter *α* + *m*_*k*_ and scale parameter 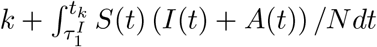.

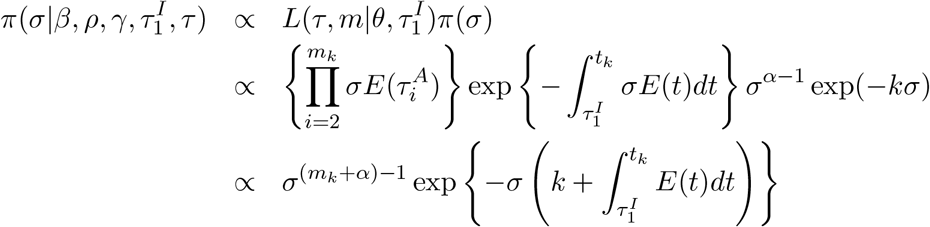

Hence the posterior distribution for *σ* is gamma with shape parameter *α* + *m*_*k*_ and scale parameter 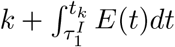.

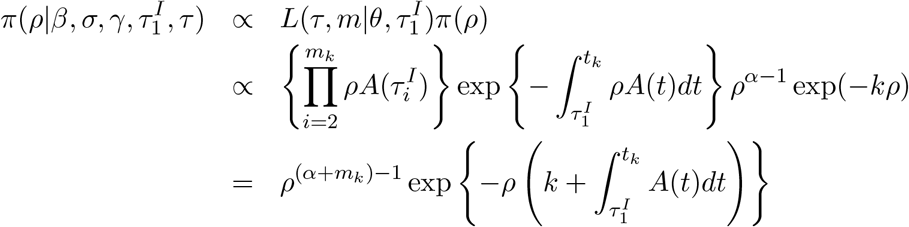

Hence the posterior distribution for *σ* is gamma with shape parameter *α* + *m*_*k*_ and scale parameter 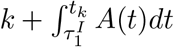.

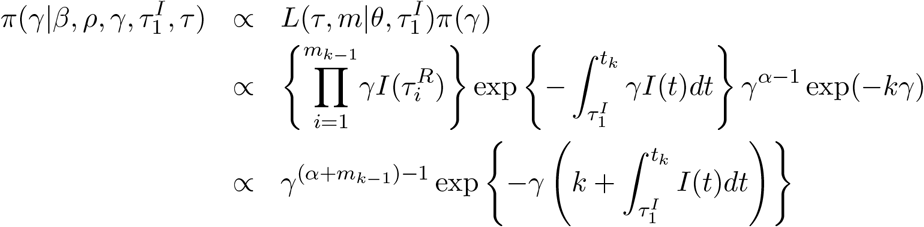

Hence the posterior distribution for *γ* is gamma with shape parameter *α* + *m*_*k*_ and scale parameter 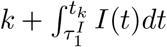.

Lastly, we calculate the posterior distribution of 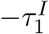, with a prior distribution of exponential, rate one.

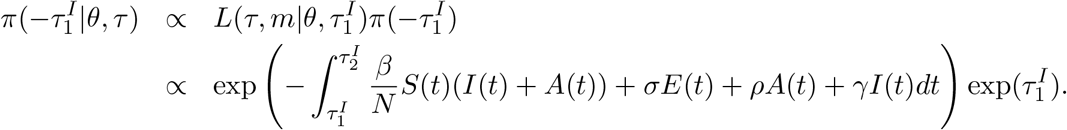

For 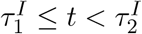, we have *S*(*t*) = *N, I*(*t*) = 1, (*E*)(*t*),*A*(*t*) = 0, and hanec

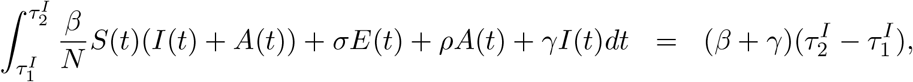

from which it follows that

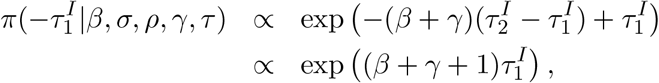

and hence the posterior of 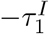 is exponential with rate *β* + *γ* + 1. Note that this formula is the same for all models.

### A.5 Symmetric Proposal

We need to show that *g*(*τ* |*τ*_*l*_)*/g*(*τ*_*l*_|*τ*) = 1. To do this, we use the fact that *g*(*τ* |*τ*_*l*_) does not depend on *τ*_*l*_. Moreover, *g*(*τ*) is a product of uniform distributions, so *g*(*τ*) also does not depend on *τ* . Therefore *g*(*τ* |*τ*_*l*_) = *c* for some constant *c*, and hence *g*(*τ* |*τ*_*l*_) = *g*(*τ*_*l*_|*τ*) = 1.

### A.6 Sensitivity to Prior

As mentioned previously, the joint prior distribution of the unknown rate parameters *θ* is made up of independent gamma distributions given by Γ(*α, k*) with mean *k/α*. In the main text, we assume that *α* is the same for the parameters *β, σ, ρ, γ*, while *k* varies and if appropriate will be denoted by *k*_*β*_, *k*_*σ*_, *k*_*ρ*_, *k*_*γ*_. In the simulations we took these to be *α* = 1 and *k*_*β*_ = *k*_*σ*_ = 3, *k*_*ρ*_ = 2, *k*_*γ*_ = 5. The prior distribution on 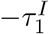is exponential with rate one, and this is independent from the *θ* vector. In Figure 9, we compare the results in the main text with results repeating the method with a different prior distribution for the SIR/SEIR/SEAIR data assuming SIR/SEIR/SEAIR models respectively. The modified prior for the comparison is *k*_*β*_ = 9*/*4, *k*_*γ*_ = 3. These were chosen as alternative reasonable parameters for the flu. The plots show that there was very little change between the two versions.

**Figure 9:**
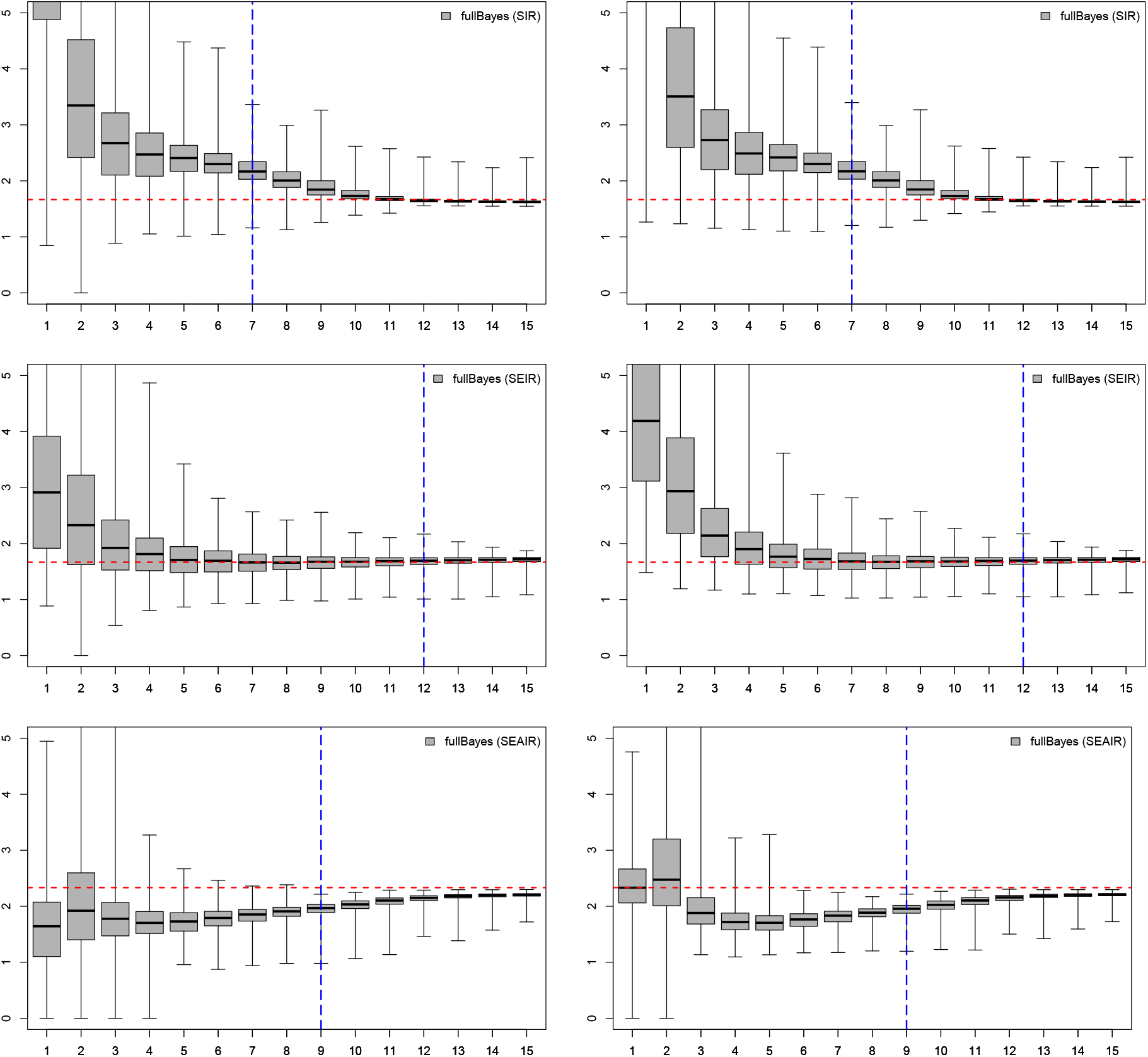
Comparison of the fullBayes method for SIR, SEIR, and SEAIR data with two different prior distributions: same as main text is on the right and the modified version is on the left. The inflection point for the epidemic is marked in blue, and the true *R*_0_ for the data is marked as a horizontal red line.

## Notes

### Competing Interest Statement

The authors have declared no competing interest.

### Funding Statement

This study was funded by the National Science and Engineering Research Council of Canada (NSERC).

### Author Declarations

No oversight body approval is needed as this is a statistics study applied to computer generated data

